# Measles Virus Genotype A in Canada’s Capital Region Wastewater Associated with Public Health Vaccination Initiatives

**DOI:** 10.1101/2024.10.11.24315327

**Authors:** Emma Tomalty, Élisabeth Mercier, Lakshmi Pisharody, Tram Nguyen, Xin Tian, Md Pervez Kabir, Chandler Wong, Felix Addo, Nada Hegazy, Elizabeth Renouf, Shen Wan, Robert Delatolla

**Affiliations:** University of Ottawa

## Abstract

The recent global resurgence of measles in 2023-2024, despite its preventability through vaccination, is a significant public health concern largely driven by decreased vaccination coverage during the SARS-CoV-2 pandemic. To address this resurgence and to restore vaccine coverage disrupted by the pandemic, Ottawa Public Health intensified vaccination efforts in 2023 and 2024. Additionally, a research initiative began in April 2024 to monitor Ottawa wastewater for measles virus (MeV) using established wastewater and environmental surveillance (WES) protocols. Given the absence of active measles cases in the Ottawa region, unexpected positive MeV detections through RT-qPCR prompted genotypic analysis as well as retrospective analysis of archived RNA samples dating back to 2020. The genotypic analysis identified positive detection to belong to genotype A, the progenitor strain of the viral vaccines, marking the first report of MeV RNA and MeV vaccine shedding in North American wastewater. Positive detections in both real-time and retrospectively analysed samples coincided with the increased vaccination efforts by Ottawa Public Health. These finding emphasize the importance of integrating genotypic analysis into WES practices to mitigate possible confounding factors, such as vaccine shedding into wastewater. Additionally, this research highlights the potential application of MeV WES for monitoring community immunization efforts in real time. Implementing the findings of this study for MeV WES, as well as for other re-emerging viruses, will enhance the accuracy of public health response and optimize resource allocation.

## 1. Introduction

Measles, a viral infection primarily affecting children, results from an infection by the measles virus (MeV) of the *Morbillivirus* genus ^1^. According to the World Health Organization (WHO), wildtype measles can be found classified into 24 genotypes distributed across 8 clades (A-H), with 3 genotypes (B3, D8, and H1) currently circulating globally and all vaccine strains belonging to genotype A ^2–4^. The WHO framework for verifying endemic measles elimination emphasizes the necessity of genotyping clinically confirmed measles cases to distinguish between measles illness from a circulating wildtype strain and a vaccine-associated illness, enabling the tracking of transmission pathways ^5^.

Despite the high contagiousness of measles infections, measles is preventable through vaccination, necessitating widespread immunity (>95%) to halt transmission. In Canada, the vaccines for measles/mumps/rubella/varicella (MMR & MMR-Var) are included in routine childhood immunization programs ^6^. However, during the pandemic, disruptions in vaccination schedules led to a concerning surge in infections. In 2022, compared to the previous year, there was an 18% rise in estimated measles cases and a 43% increase in measles-related deaths globally ^2,3,7^. While Canada maintains no ongoing transmission of MeV, the risk persists due to incoming travelers from regions where the virus is prevalent. Consequently, susceptible populations, such as under-immunized communities or individuals with fading immunity, remain vulnerable to introduced infections ^8^.

For over a decade, wastewater and environmental surveillance (WES) has been used to monitor community transmission and infection rates of non-enveloped viruses spread via the fecal-oral route, such as poliovirus, hepatitis virus, and norovirus ^9,10^. During the SARS-CoV-2 pandemic, the full potential of WES was realized and is now used worldwide to track various respiratory viruses, including SARS-CoV-2, influenza A, influenza B, and respiratory syncytial virus. MeV, another respiratory virus, is reported to be primarily detected in the urine of infected individuals, enabling its detection in wastewater ^11,12^. Recent studies have demonstrated the use of WES as an unbiased surveillance method for MeV through detection of wildtype strains in wastewater during measles outbreaks in affected communities ^13–15^.

Following the global increase in measles cases post-pandemic, Ottawa Public Health intensified its vaccination efforts in 2023 and 2024 to restore vaccine coverage lost during the pandemic years of 2020 to 2022. To complement these efforts and address rising concerns about measles, a research initiative was launched for measles surveillance in wastewater, utilizing established WES systems used for monitoring respiratory viruses. The detection of an unexpected MeV signal in Ottawa’s wastewater, concerning due to the absence of known active measles cases in the city, prompted a genotypic investigation to identify the specific strains present. Analysis revealed that the detected MeV was not wildtype but rather vaccine type, marking it the first report of MeV RNA and MeV vaccine shedding in North American wastewater and second report of MeV vaccine shedding in the world ^15^. This study highlights the critical insight of MeV vaccine shedding into wastewater as a confounding factor and underscores the necessity of genotyping within WES practices to distinguish between vaccine type and wildtype viruses. By ensuring accurate differentiation, public health responses can be appropriately guided, avoiding misinterpretations and enabling precise actions based on the actual measles activity in the community.

## 2. Materials and Methods

### 2.1 Wastewater Sampling, Sample Nucleic Acid Extraction, and RT-qPCR Analysis

Twenty-four-hour composite primary clarified sludge samples were collected from the City of Ottawa’s only water resource recovery facility as part of an ongoing wastewater surveillance program. Daily samples between April 6, and May 31, 2024, were processed for nucleic acid extraction within 48 hours of arrival at the laboratory using previously described methods ^16^. Retrospective analysis of archived extracted RNA from wastewater samples collected from July 2020 to March 31, 2024 were analysed by RT-qPCR after being stored at -80°C for periods ranging from 1 to 45 months. Frequency of samples tested for retrospective analysis is detailed in Table S.1. Viral MeV was quantified using the assay described by Hummel et al. (2006), targeting the nucleoprotein ^17^. Additional details for primers, probes and cycling conditions are summarized in supplementary information and Table S.2.

### 2.3 Sample Amplification, Purification, Sanger Sequencing and Strain Identification

Wastewater samples showing a positive MeV detection through RT-qPCR were further analysed to identify the strain circulating in the community. For these samples, a nested-PCR approach previously described by Brzovic et al. (2022), was used to increase the sensitivity; two sequential amplification reactions were performed using two different sets of primers ^18^. A description of the primers and cycling conditions can be found in supplementary information and Table S.2. The samples were then sequenced by Sanger sequencing of the N450 nucleotide region according to the WHO guidelines at the Ottawa Hospital’s Research Institute StemCore Sequencing Facility using primers listed in Table S.3 ^19^. The sequences were compiled and edited using Snapgene Viewer version 7.2.0. Genotype and clade were determined by comparing the sequences with the NCBI Blast database of sequences and phylogenetic analysis using Genome Detective.

## 3. Results and Discussion

### 3.1 MeV Vaccine Shedding: Detection of Measles Genotype A

Positive MeV signal was detected in 8% of wastewater samples (11/135) (Figure 1). The first positive detection was on August 4, 2023 and was the only detection in 2023. All subsequent positive detections were in 2024. Specifically, nine of the positive detections were from the daily samples and only two positive detections were from the retrospective analysis of archived RNA samples.

**Figure 1.**
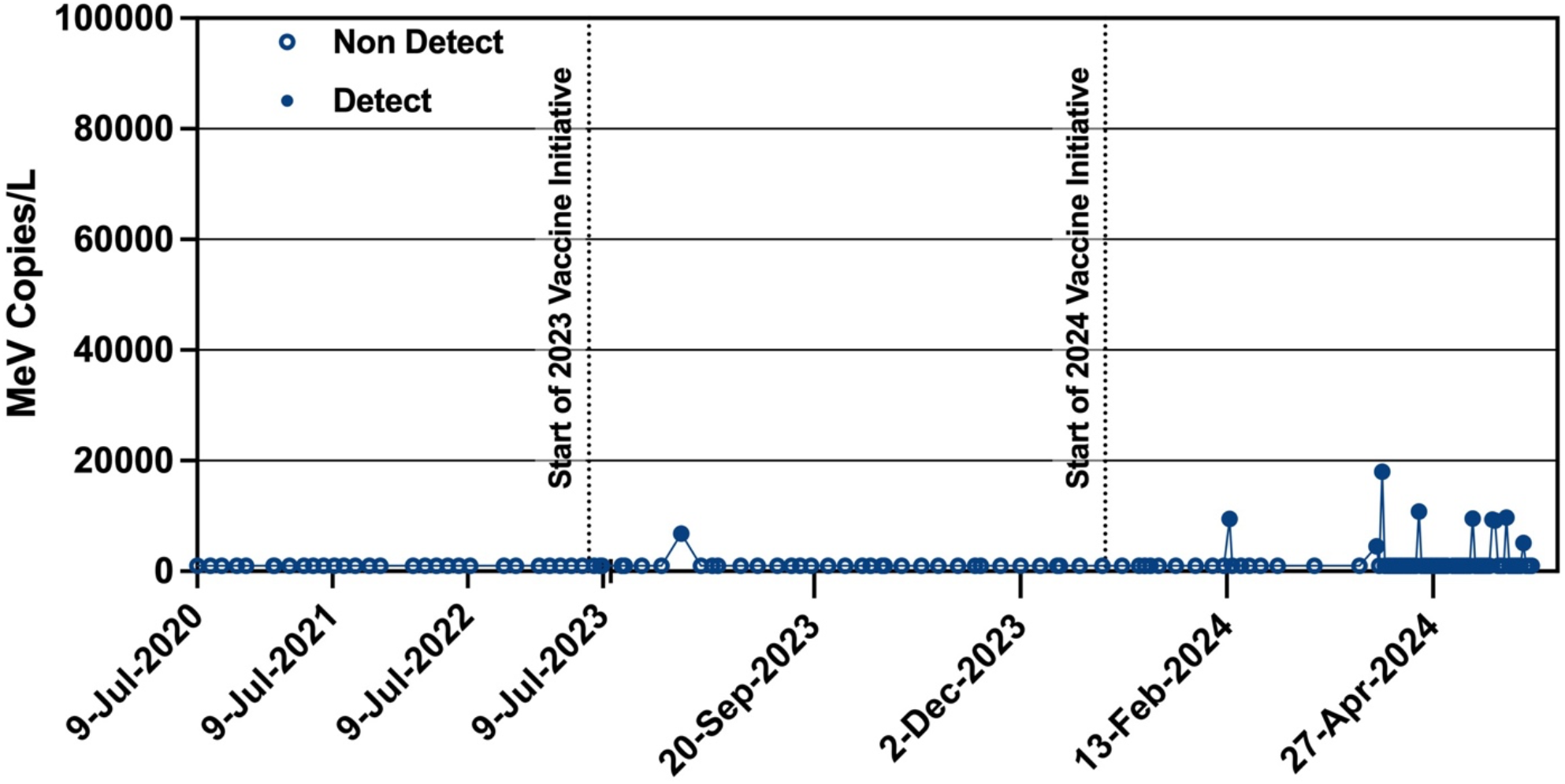
Genome copies of MeV per litre of wastewater sample from July 2020 to May 31, 2024. The open circles indicate non detects and closed circles indicate detects. The dashed lines indicate the approximate start of increased vaccination efforts by Ottawa Public Health in 2023 and in 2024.

The detection of MeV in wastewater was a public health concern in the city of Ottawa, particularly as there were no known active measles cases in the population at the time ^20^. As such, in accordance with WHO framework, genetic characterization of MeV upon detection in wastewater was carried out to monitor virus circulation and to distinguish the virulent strain ^21,22^. Two of the wastewater samples with positive MeV detection were Sanger sequenced for genomic analysis: wastewater samples collected on April 6, 2024 and May 22, 2024. Sequence comparison with NCBI Blast database revealed a 100% homology with the Moraten vaccine strain (NCBI Accension # AF266287) and the Schwarz vaccine strain (NCBI Accension # AB591381). Phylogenetic analysis using the measles typing tool on the Genome Detective website confirmed identification of measles genotype A (Figure S.1). According to WHO, all wildtype strains of genotype A are extinct, indicating that the detected MeV was the vaccine strain ^23^. This study represents the first detection of MeV vaccine in North American wastewater, indicating vaccine-shedding of live-attenuated MeV. This outcome was unexpected, as most reports of MeV detections in other regions involved circulating wildtype strains ^13,14^.

It is noted that low recovery of MeV from RNA samples stored at -80°C, from July 2020 to March 31 2024, in this study may be presented as non-detects due to viral loss during storage. Samples collected between April 6, and May 31, 2024, were not archived and instead were processed and analyzed within 48 hours of collection. Research on the effects of storage at -80°C of SARS-CoV-2 RNA from wastewater samples found no significant difference in recovery when re-quantified after 16 months storage ^24^. However, the effects of storage on MeV RNA from wastewater has not yet been studied.

Additionally, the viral signal from fresh samples was already low, consequently it is stipulated that even a small amount of decay could lead to loss of viral signal in the archived samples. Therefore, future work should be done to understand the stability of MeV RNA in wastewater.

### 3.2 Public Health Measles Vaccine Initiatives Associated with MeV Measurements in Wastewater

The measles vaccines currently available in Ontario, Canada, and hence in the city of Ottawa, are Priorix (MMR), Proquad (MMR-Var), and MMR II (MMR). MMR II and Proquad are manufactured by Merck & Co., using the Moraten strain ^25^. Priorix is manufactured by GlaxoSmithKline Biologicals using the Schwarz strain ^26^. Both the Moraten and Schwarz strain were developed from the Edmonston isolate and previous analysis of the sequences of the Moraten and Schwarz strains indicated identical coding sequences belonging to MeV genotype A ^27,28^. Research to-date indicates that vaccine shedding occurs with vaccines containing replicative viruses, such as live-attenuated viral vaccines ^29^. This phenomenon has been demonstrated through the detection of live-attenuated viral vaccines for polio and rotavirus in wastewater ^30,31^. Previous studies examining the shedding and persistence of MeV in vaccinated individuals has demonstrated the presence of viral RNA in urine samples 1 to 15 days post vaccination with the possibility of slow viral clearance from the body persisting for up to 100 days post vaccination ^32–34^. Due to the strict vaccine storage and handling guidelines in Canada, the viral signal identified in the samples analysed in this study is unlikely to be due to disposal down the drain but instead is stipulated to originate from individuals recently vaccinated, as well as those who continue to shed the vaccine over extended durations ^35^.

Given that 91% (10/11) of MeV detections occurred in 2024, with no signals detected in wastewater samples prior to 2023, the authors investigated vaccination data to identify factors potentially contributing to this temporal pattern. Reduced vaccination rates during the pandemic in combination with a global rise in measles cases and reduced travel restrictions has led to an increase in the number of measles cases seen in Ontario ^20^. As of May 31, 2024, there were 23 confirmed cases of measles reported in Ontario occurring in nine different public health units; no measles cases were reported in Ottawa to date. In an effort to restore vaccine coverage post-pandemic and in response to increased measles activity in Ontario, Ottawa Public Health increased vaccination initiatives by sending automated messages via text, email or phone call to households with one or more children overdue for vaccinations. These messages, sent periodically between June and October 2023 and January and March 2024, were intended to drive families to the school clinics provided by Ottawa Public Health’s Immunization Program or to primary care providers. Additionally, due to increased risk of exposure, health care workers often receive routine immunization booster shots, including MMR/MMR-Var vaccines ^36^.

Vaccination data provided by Ottawa Public Health indicated a rise in vaccines administered in 2023 compared to previous years (2020 – 2022) (Figure 2). There was a notable delay in reporting vaccine administrations during and after the SARS-CoV-2 pandemic years. For instance, on average, only about 60% of vaccinations were reported within the same year they were administered, with remaining administrations being reported periodically in subsequent years (Figure S.2). Consequently, Ottawa Public Health may not have true, up-to-date numbers of total vaccines administered. To address the discrepancy between the date of administration and the date of reporting, the rate of vaccine administrations was calculated via the mean and standard deviation of administered vaccines that were reported the same month they were administered. This approach helps mitigate biases within the dataset and enables a more accurate comparison of the vaccination rates from 2020 to 2024. A caveat within the dataset received is that vaccine reporting is lower than usual therefore the rate of vaccine administrations may be higher than is visible in the reported data. A paired t-test with significant alpha of 0.05, revealed that vaccination rates were significantly higher in 2023 compared to 2020, 2021, and 2022 (p = 5.5E-06, p = 2.2E-09, and p = 1.1E-08, respectively). This increase corresponds with the increase in vaccine initiatives by Ottawa Public Health through automated messages. Vaccination rates in 2024 were also significantly higher than 2020, 2021 and 2022 (p = 2.5E-03, p = 2.5E-03, p = 3.6E-03, respectively). Though there was no significant increase between 2023 and 2024 (p = 0.17), this is likely due to carry over of increased vaccine initiatives starting in 2023. The increase in vaccination rates in 2023 and 2024 aligns with the detection of MeV vaccine shedding in wastewater during the same period, reinforcing the link between heightened vaccination efforts and the observed signals (Figure 2). The minimal detections from archived wastewater RNA collected in 2023 is stipulated to be due to viral degradation during extended storage.

**Figure 2.**
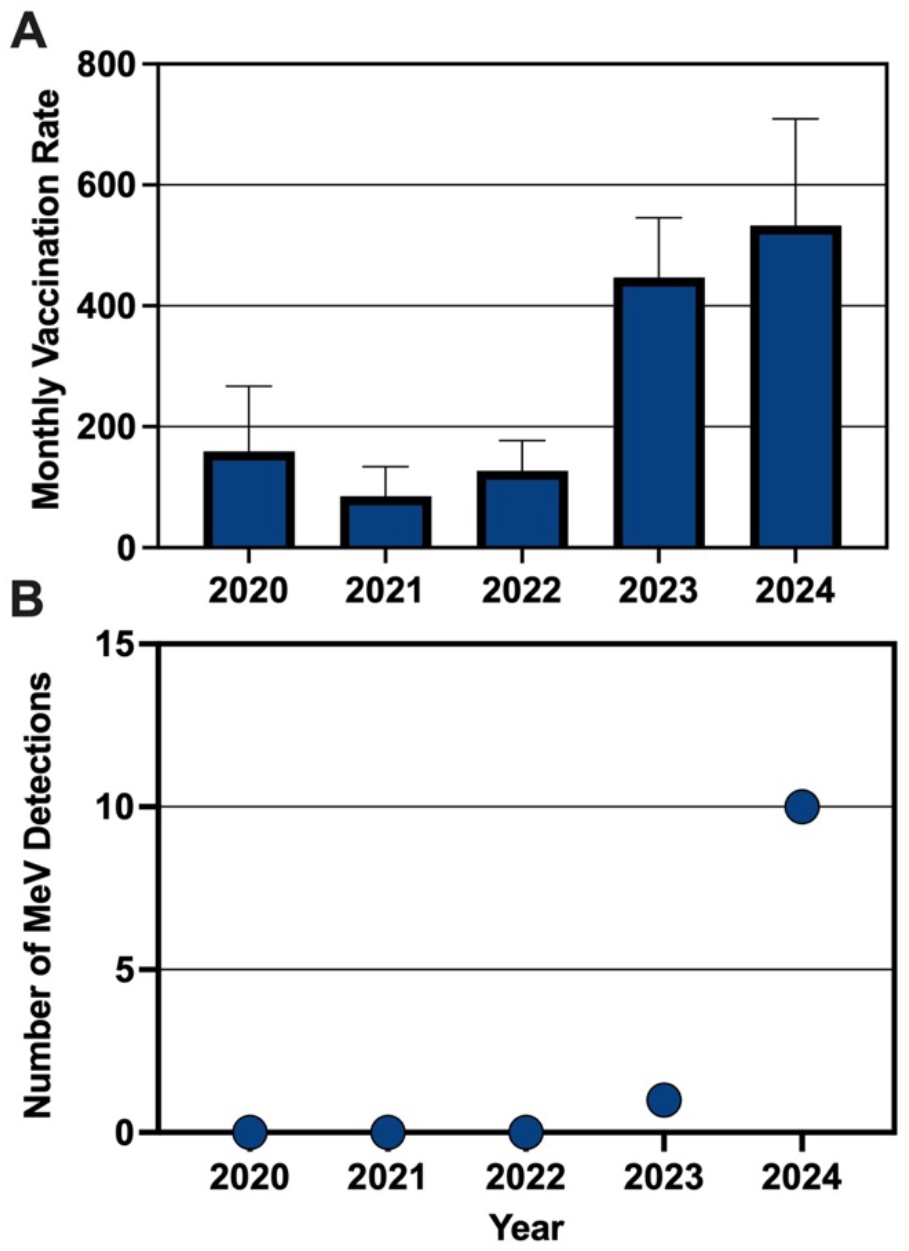
**A**. Monthly vaccination rate during the SARS-CoV-2 pandemic shown as the average number of reported vaccines administered per month from April 2020 to June 2024. **B**. Increases in monthly vaccination rates in 2023 and 2024 occur during the same periods of MeV vaccine strain detection in wastewater.

### 3.3 Measles WES and Public Health Applications

WES has the potential to serve as an important public health tool by detecting MeV RNA in wastewater and hence identifying risk of measles transmission prior to clinical cases presenting within a community. The time from exposure to rash onset of clinical measles cases averages 14 days with patients considered infectious during the prodromal phase approximately four days before rash onset ^23^. In Ontario individuals presenting with fever, maculopapular rash for at least three days, and at least one of: cough, coryza or conjunctivitis are suspected measles cases requiring confirmation via virus isolation (culture) or RNA detection (PCR) from a collected clinical specimen ^37^. Only measles cases which meet criteria for confirmed case classification according to the Ontario Ministry of Health and Long-Term Care surveillance are included in reported case counts. Mild and asymptomatic cases of measles may not meet criteria and therefore may not be reported while detection of MeV in urine samples from asymptomatic individuals has been demonstrated in literature ^38^. One of the major and enhanced applications of measles WES is to identify outbreak hotspots in the community, thereby facilitating prompt decisions to control and mitigate circulation and transmission of MeV ^14,39^. In the event of a detected measles signal in wastewaters, public health authorities would swiftly mobilize resources, initiating urgent clinical measures, including enhanced surveillance and rapid laboratory confirmation via PCR or serology, leading to promp isolation of suspected cases and immediate contact tracing ^37^. These actions could be accompanied by expedited vaccination efforts to prevent further transmission due to the highly contagious nature of measles and the risk of large-scale outbreaks ^40^. However, accurately interpreting a MeV signal in wastewater requires identifying the strain because vaccination rates and recent immunization efforts can confound results. As such, routine genotypic analysis of MeV in wastewater is essential to ensure that signals from the vaccine strain are correctly identified, allowing public health resources to be directed appropriately.

Hence, the detection of MeV signal in wastewater offers valuable insights beyond being an early warning system of measles re-emergence in a community, it can also track vaccination coverage within communities. Similar applications have been demonstrated for the live-attenuated vaccine-virus for Polio in Sweden, Finland, and Cuba following a vaccine campaign ^30,41^. In Canada, maintaining the elimination of endemic measles transmission relies heavily on strategies such as effective surveillance and high immunization coverage. Continual monitoring of Ottawa wastewater for MeV provides valuable insights, allowing public health officials to track re-emergence and the impact of vaccination campaigns in real-time, thus eliminating the wait for reported administrations. Given the pattern of delays in reported administrations, the clinical data would likely be outdated and may not provide the most accurate or relevant information in light of increased vaccination efforts.

This study marks a significant milestone as the first to detect and identify the vaccine strain of MeV in North American wastewater and compliments the findings of MeV vaccine strain in South Africa ^15^. It underscores the need for routine genotypicc analysis to prevent the misallocation of public health resources in response to MeV WES signals that do not pose a transmission risk. Additionally, this research highlights the potential application of MeV WES for tracking community immunization efforts in real time. The findings of this study highlights the potential of WES for other re-emergent infections to enhance outbreak preparedness, prevention, and response.

## Data Availability

All data produced in the present study are available upon reasonable request to the authors

## Acknowledgements

The authors wish to acknowledge the help and assistance of the the City of Ottawa, Ottawa Public Health, Public Health Ontario and all their employees involved in the project during this study. Their time, facilities, resources and thoughts provided throughout the study helped the authors greatly. The authors also wish to specifically outline the assistance of the Ottawa Public Health Epidemiology and Evidence team. The authors would like to acknowledge the assistance of Dr. Alessandro Zulli from the Department of Civil and Environmental Engineering, Stanford University. The authors would like to acknowledge funding from the CIHR Applied Public Health Chair in Environment, Climate Change and One Health, awarded to Robert Delatolla.

