## Supplementary material for "Measles Virus Genotype A in Canada’s Capital Region Wastewater Associated with Public Health Vaccination Initiatives": Table S.1 Table S.2 Table S.3 Figure S.1 Figure S.2

**Table S.1:** Frequency of retrospective analysis of archived wastewater RNA from July 2020 to March 2024. Missing dates were due to lack of archived RNA.

| Year | Number of Samples Analysed | Frequency |
|---|---|---|
| 2020 | 5 | starting July, once per month, missing December |
| 2021 | 11 | Once per month, missing December |
| 2022 | 8 | Once per month, missing January, August, September and December |
| 2023 | 40 | Once per month January to June. Starting July, once per week |
| 2024 | 17 | Once per week, ending March |

**Additional details for RT-qPCR analysis**

All RT-qPCR reactions were prepared using TaqMan™ Fast Step Master Mix (Thermo Scientific) according to the manufacturer's guidelines for a final reaction volume of 10 µl and quantified using a thermal cycler (CFX96, Biorad). Samples from April 6 to May 31, 2024, were run in eight technical replicates, while retrospective samples were run in triplicate, each with non-template controls and a five-point standard curve usinga MeV G-block (IDT) [1]. The assays limit of detection (ALOD) and limit of quantification (ALOQ) were approximately 3.66 and 3.74 copies/reaction respectively following recommended MIQE guidelines [2]. For all qPCR analyses efficiency ranged from 90 – 110%, $R^2$ values were greater than 0.95 and MeV was considered positive if at least two wells with Ct < 40.

**Table S.2:** Amplification target regions, primers and cycling conditions

| Amplicon length | Primers/Probes | Cycling Conditions | Reference |
|---|---|---|---|
| 75 bp | MVN 1139F 5' TGGCATCTGAACTCGGTATCAC 3'<br>MVN 1213R 5' TGTCCTCAGTAGTATCGATTGCAA 3'<br>MVNP 1136 5' FAM-CCCCGAGGATGCAAGGCTTGTTTCAGA-BHQ1 3' | **RT:** 50°C for 5 min (1 cycle)<br>**RT Inactivation/ Initial Denaturation:** 95°C for 20s (1 cycle)<br>**Denature:** 95°C for 30s (44 cycles)<br>**Anneal/Extend:** 60°C for 30s (44 cycles) | Hummel et al. 2006[3]<br><br>RT-qPCR for quantification |
| 439 bp | MVN 3 5' GGATGAGGCGGACCAATACT 3'<br>MVN 6.1 5' TGACCATGCTGCCATAGCTT 3' | **Initial Denaturation:** 98°C for 30s (1 cycle)<br>**Denaturation:** 98°C for 10s (35 cycles)<br>**Annealing:** 55°C for 30s (35 cycles)<br>**Extension:** 72°C for 60s (35 cycles)<br>**Final Extension:** 72°C for 120s<br>4°C, hold | Brzovic et al., 2022[4]<br><br>Nested PCR for amplification |
| 695 bp | MVN 5 5' GGAGTAGGAGTGGAACTTG 3'<br>MVN 6 5' TCTGCCATCGGCTCCAATCG 3' | **Initial Denaturation:** 98°C for 30s (1 cycle)<br>**Denaturation:** 98°C for 10s (35 cycles)<br>**Annealing:** 55°C for 30s (35 cycles)<br>**Extension:** 72°C for 30s (35 cycles)<br>**Final Extension:** 72°C for 120s<br>4°C, hold | |

**Additional information for sample amplification and purification**

The 450 nucleotides encoding the carboxylterminal 150 amino acids of the nucleoprotein were sequenced according to the WHO guidelines as the minimum amount of sequence data required for determining the genotype of MeV [5]. Synthesis of cDNA was first performed using SuperScript™ IV First-Strand cDNA Synthesis Reaction following the manufacturer guidelines (Invitrogen). The primary and secondary PCR were prepared using Q5 Hot Start High-Fidelity 2X Master Mix (New England Biolabs) according to the manufacturer guidelines for a final reaction volume of 50 µl. The PCR product from the primary PCR and the secondary PCR were visualized through gel electrophoresis followed by gel purification using the QIAquick Gel Extraction Kit (Qiagen) as per manufacturer guidelines. The concentration of the purified amplicon product was quantified using a Nanodrop (Thermo Scientific) followed by dilution to a final concentration of 1 ng/µl.

**Table S.3:** Primers used for Sanger Sequencing

| Amplicon Length | Primers | Reference |
|---|---|---|
| 695 bp | MVN 5 5' GGAGTAGGAGTGGAACTTG 3'<br>MVN 6 5' TCTGCCATCGGCTCCAATCG 3' | Brzovic et al., 2022[4] |
| 634 bp | MeV 216 5' TTGAGCTATGCCATGGGAGT 3'<br>MeV 214 5' TAACAATGATGGAGGGTAGG 3' | CDC, 2018[5] |

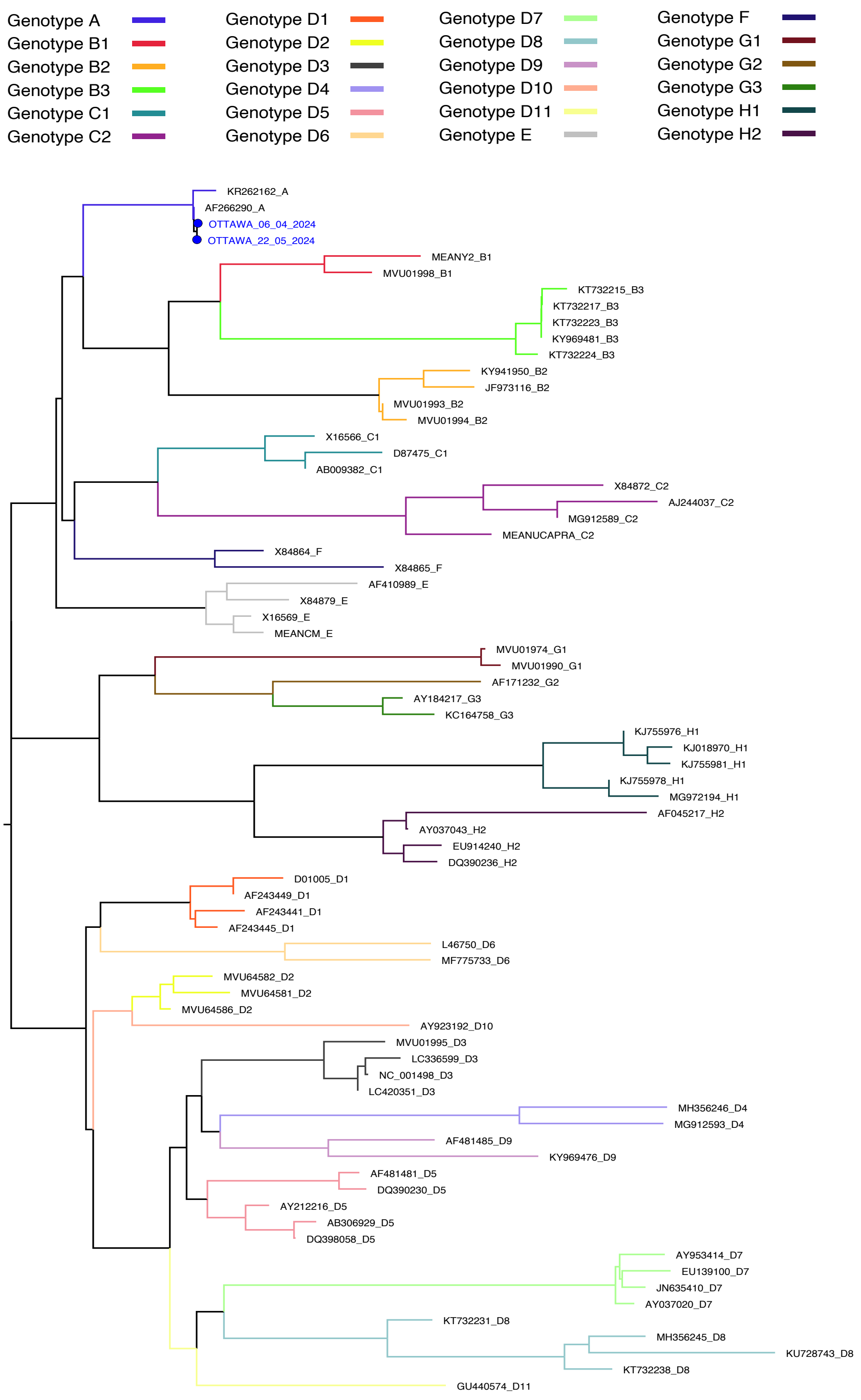


**Figure S.1:** Phylogenetic analysis of Sanger Sequencing results using the measles typing tool on the Genome Detective website confirmed identification of genotype A indicating MeV vaccine strain.

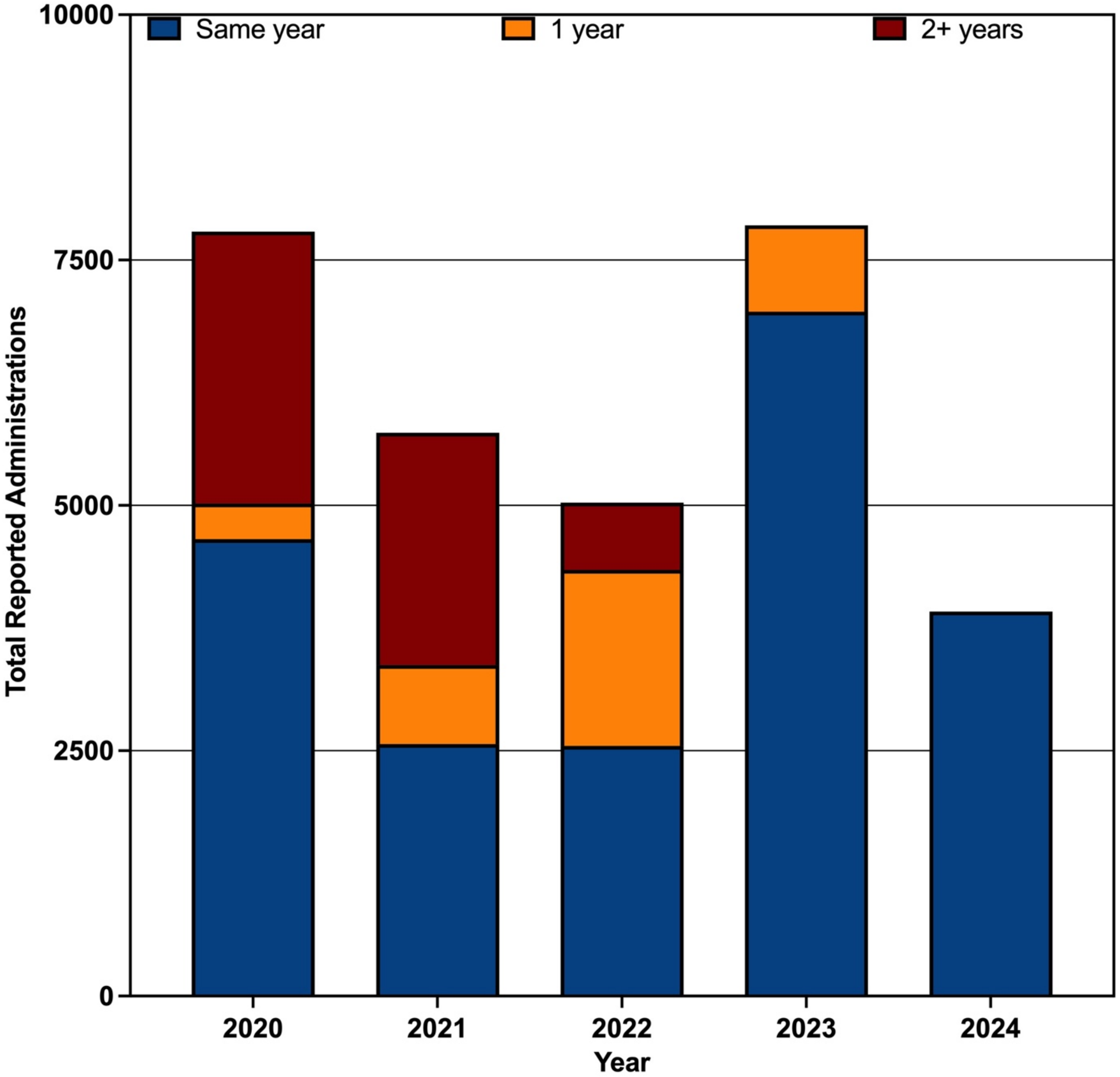


**Figure S.2:** A bar graph showing the delay in reporting vaccine administrations. The bar graph is comprised of three sections: blue represents the vaccinations reported within the same year, orange represents vaccinations reported the next year, and red represents vaccinations reported two or more years after administration. The total reported vaccines for 2020 to 2023 are from January to December, while total reported vaccines for 2024 are from January to June.

<u>**References**</u>


1. Mercier, E. *et al.* Municipal and neighbourhood level wastewater surveillance and subtyping of an influenza virus outbreak. *Sci Rep* **12**, (2022).

2. Bustin, S. A. *et al.* The MIQE Guidelines: Minimum Information for Publication of Quantitative Real-Time PCR Experiments. (2009) doi:10.1373/clinchem.2008.112797.

3. Hummel, K. B., Lowe, L., Bellini, W. J. & Rota, P. A. Development of quantitative gene-specific real-time RT-PCR assays for the detection of measles virus in clinical specimens. *J Virol Methods* **132**, 166–173 (2006).

4. Brzovic, M. *et al.* Measles cases in Split-Dalmatia County (a Croatian tourist region), in May–July 2019: outbreak report and lessons learnt. *Eur J Public Health* **32**, 948–954 (2022).

5. CDC. annex-c1-measles-genotyping-rt-pcr.
